# Spectrally and temporally segmented regression of nuisance signals in high-speed resting-state fMRI

**DOI:** 10.64898/2025.11.26.25341017

**Authors:** Khaled Talaat, Bruno Sa de La Rocque Guimaraes, Stefan Posse

## Abstract

**Purpose:** Prior work has shown that whole-band linear regression of nuisance signals can introduce artifactual connectivity in high-frequency resting-state fMRI. Errors of motion regressors and non-stationarity of nuisance signals exacerbate artifacts. Here, we introduce spectral-temporal segmentation of regression vectors to decouple regression in different frequency bands to reduce motion artifacts.

**Methods:** An alternative approach to whole-band linear nuisance regression is introduced in the present work relying on spectral segmentation of the motion parameters into k-bands using non-causal or FIR filters, with whole-band regression of the filtering residual, and temporal segmentation of regression vectors. The methodology was tested in computer simulations and in-vivo data. Resting-state networks in five healthy controls and two brain tumor patients using high-speed fMRI (TR >= 205 ms) were mapped using the present approach combined with spectrally constrained regression of physiological noise and the results were compared to the conventional whole band regression approach.

**Results:** Computer simulations showed high tolerance to frequency dependent errors in regression vectors. Motion and physiological noise artifacts in-vivo were substantially reduced without introducing artifactual connectivity. Artifactual connectivity decreased asymptotically with increasing number of frequency bands without decreasing connectivity in major resting-state networks. Connectivity above 0.3 Hz in-vivo was consistent with that in traditional low-frequency networks.

**Conclusions:** Spectral-temporal segmentation of regression vectors is a powerful approach to reduce artifacts from non-stationary high-bandwidth nuisance signals.

## INTRODUCTION

Resting-state functional MRI (rsfMRI) has emerged as an adjunct to task-based fMRI, allowing for simultaneous mapping of functional connectivity across dozens of resting-state networks (RSNs) to study a wide range of neuroscience and clinical applications with more than one hundred resting-state networks identified in group studies and interindividual differences in network topology detectable in individual subjects [1–8]. Advances in high-speed fMRI [9–16], which enable unaliased sampling of physiological signal fluctuations, have increased sensitivity for mapping functional connectivity and detecting dynamic changes [11, 14, 16] compared with conventional echo-planar imaging (EPI) techniques. Recently, several studies using high-speed fMRI techniques have reported different potential resting-state networks (RSNs) at high frequencies (up to 5 Hz) [17–22], suggesting that functional integration between brain regions at rest occurs over broader frequency bands than previously thought [20, 22].

The majority of published studies on high-frequency resting-state fMRI (hfrsfMRI) utilized nuisance regression. Critical concerns have been raised on the use of conventional regression techniques in the post-processing of rapidly sampled fMRI data [23, 24]. In particular, it was demonstrated that the use of whole-band linear regression (i.e. using regressors that cover the entire frequency band) in motion and physiological noise correction can pass structured network patterns from the conventional low frequency band artificially to higher frequencies and introduces artifactual high frequency connectivity above those predicted from the canonical model which sets an upper limit at 0.3 Hz [23]. Resting-state fMRI is highly sensitive to movement which obscure networks as well as create false positive correlations [25–27]. Motion regression utilizing up to 24 realignment parameters [27], spike regression [27], motion scrubbing [28], PCA based regression of nuisance signals using aCompCor [29] and a range of ICA based approaches [30] have been developed to remove motion and physiological pulsation artifacts in resting-state fMRI. The recently developed Harmonic Regression with Autoregressive Noise (HRAN) method using single-frequency temporally dynamic regression [31] has been shown to remove physiological noise while leaving the neural signal intact, thereby increasing detection of task-based activation in fast fMRI. However, the applicability of these approaches to high-speed fMRI and high-frequency resting-state fMRI remains insufficiently characterized. Alternate methods such as conventional finite impulse response (FIR) filtering of physiological noise suffer from extensive data losses as large portions of the spectra in cortical areas may be contaminated with respiration, cardiac pulsation, and their harmonics, some of which may be aliased in case of long repetition times.

An alternative approach to whole-band linear nuisance regression is introduced in the present work, relying on spectrally and temporally segmented regression of nuisance regressors. Notably, the approach does not require spectral segmentation of the scan data itself. This approach allows for one-shot independent regression in different spectral bands to prevent the artifactual injection of low frequency connectivity into high frequency bands described in [23]. By calculating an independent weight for each spectral band, the present approach assigns low weights to spectral bands where the regressor and the signal exhibit minimal overlap and thus mitigates the injection of signal from uncorrelated spectral parts of the nuisance regressor into the signal. The approach is applied to motion regressors where inter- and intra-slice errors at higher frequencies, which may be introduced by rapid head movement, result in regressor errors. Regression of filtering residuals was added as an important step to compensate regression errors when using finite impulse filter-based segmentation. Physiological noise regression of respiratory and cardiac pulsation is implemented as a second step using slice-dependent phase shifts of regressors to account for regional differences in nuisance signal phase. Spectral segmentation is combined with temporal segmentation, which, as our previous work has shown, reduces artifacts in resting-state connectivity that are due to signal instability and transients ([32, 33]). Spectral segmentation is characterized in computer simulations of a 2-node resting-state network. The combined spectral and temporal segmentation approach is applied in vivo using resting-state fMRI scans acquired in five healthy controls and in two patients with brain tumors with TR as short as 205 ms. The physiological noise regression step is compared with HRAN.

## METHODS

### Spectrally and temporally segmented regression

Two consecutive procedures were developed for motion regression and physiological noise regression (**Fig. 1**). Motion regression involves construction of regression vectors from measured rigid-body translations and rotations, which are filtered in the spectral domain into k bands using either a non-causal filter or finite impulse response (FIR) filter. The non-causal filter is implemented by applying an FFT to the regression vector time course, zeroing of the spectral data outside of the segment of interest and then applying an inverse FFT. Discrepancies between the sum of the spectral segments and the original regressor arise at the interfaces of spectral segments, in particular for an FIR filter. Residuals can also arise in non-causal filtering if there are overlapping bins between the spectral segments which can be mitigated by using non-overlapping bands or by filtering the residual. The residual from filtering is obtained by regressing the original whole-band regressor from the sum of the spectral bands. This residual is then included as an additional regression vector. A constant vector is also included as described in Gembris et al. [34]. Any zero vectors that might arise during the spectral segmentation or residual calculation process must be excluded from the regression matrix. The subsequent steps involve temporal segmentation of the data and regression vectors into *n* segments, calculation of regression coefficients, and subtraction of the regression series from the original signal, 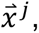 to obtain the corrected signal, 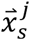 as expressed in Equation 5.

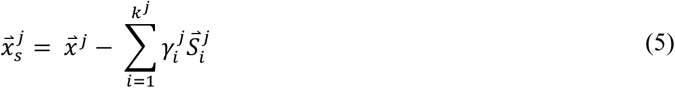

where *j* is the index for the temporal segment, 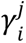 is a calculated weighting coefficient, 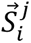 are the regression vectors and *k^j^* is the number of regression vectors within segment *j*; or in other words *k^j^* is the number of spectral segments in temporal band *j*. The corrected signal in a voxel is the concatenation of 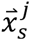 in time as expressed in Equation 6.

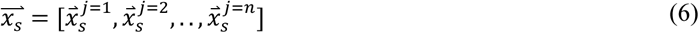

**Figure 1:**
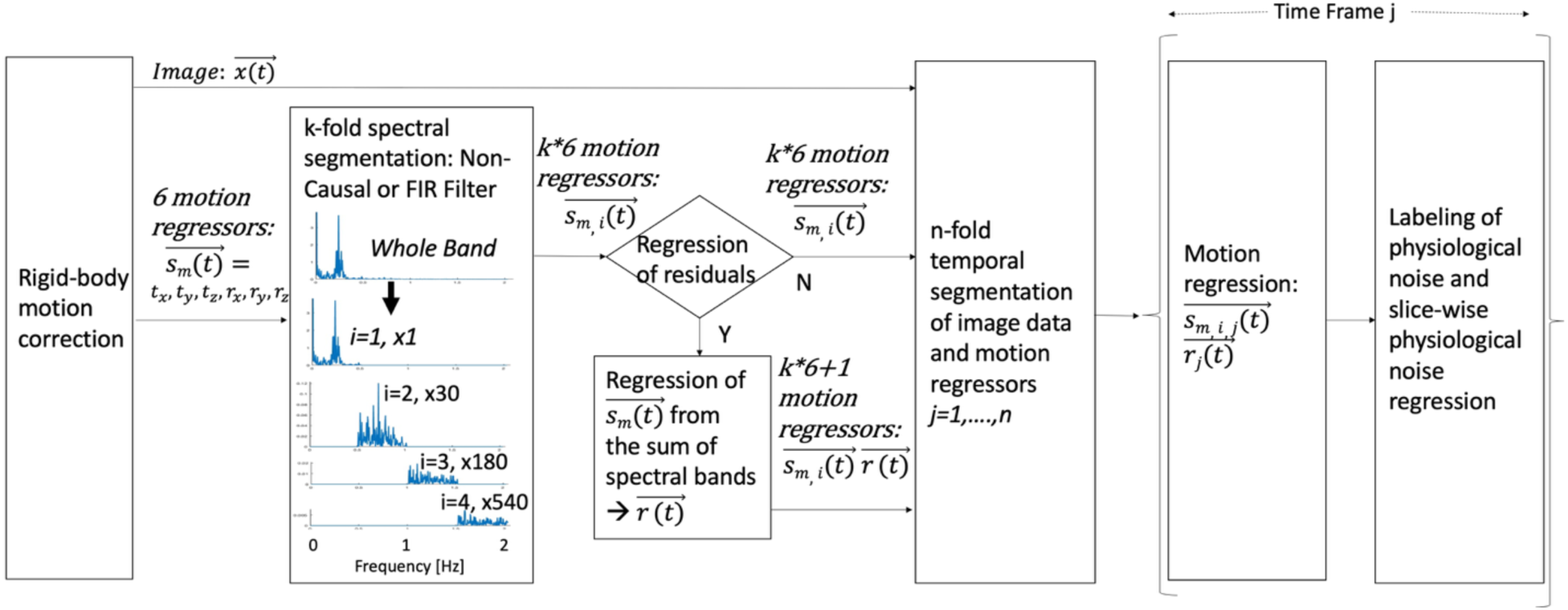
Signal processing pipeline of spectrally segmented regression of motion parameters and slice-specific regression of physiological noise within temporal windows. The example of a motion parameters spectrum which is segmented into 4 spectral bands shows a prominent peak at the respiratory frequency.

The coefficients of regression can be obtained by minimizing the quadratic form 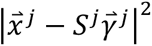 as described by Gembris et al. [34]. This minimization results in Equation 7.

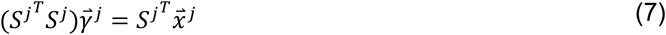

Physiological noise regression involves manual labeling of spatially averaged physiological noise within cerebral-spinal fluid (CSF) and white matter (WM) in the frequency domain (i.e. selection of upper and lower frequency bounds in reference to the range of respiratory and cardiac rates that were measured on the scanner) in each of the time segments, creation of spatial masks for each of the labeled frequency bands based on a power-spectral integral threshold relative to a labeled frequency range that is free of physiological noise (threshold: 3.0), spatial averaging of physiological noise signals within the masks and creation of a set of physiological noise regressors for each frequency band and temporal segment. Multiple frequency bands may be selected to represent physiological noise features. The regression vectors for physiological noise are subsequently time shifted in each slice separately to account for inter-slice time delays in physiological signal pulsation and optimal time delays are found by minimizing the power-spectral integral of the labeled feature after regression over entire slices. Iterative minimization is used to search for optimal time shifts with a user-defined maximum expected time shift between any two slices in the brain, which was empirically determined to be less than 3 TRs for the data sets that were investigated (see below).

Regression of motion and physiological noise is performed in temporal windows whose minimal length is constrained by the bandpass characteristics of sliding-window regression, i.e. the low frequency cut-off described in our recent work [32]. In that study, we demonstrated effective nuisance regression using window widths between 8 and 30 s. Larger window widths increase the sensitivity to instabilities of cardiac and respiratory pulsation rates which broaden labeled frequency bands.

The method was implemented in a custom MATLAB tool (TurboFilt) that features functions for seed selection, spatial mask generation and application, non-causal and FIR frequency filtering, time-course extraction and frequency analysis, spectral feature mapping, seed-based correlation analysis, and signal simulation.

### Simulations

Simulations were performed to verify that the proposed method mitigates the injection of artifactual correlations into higher frequencies and effectively recovers the original signals prior to injection of noise.

In a first step, we applied whole-band and frequency segmented regression with two frequency bands with 0.25 Hz bandwidth each to the simple case described in Figure 1 of Chen et al. [23] where the measurement vector and the nuisance regressor both contain low (0.04 Hz) and high (0.4 Hz)-frequency components (**Fig.2**). Non-causal filtering was used to obtain the two segments (below 0.25 Hz and above 0.25 Hz).

**Figure 2:**
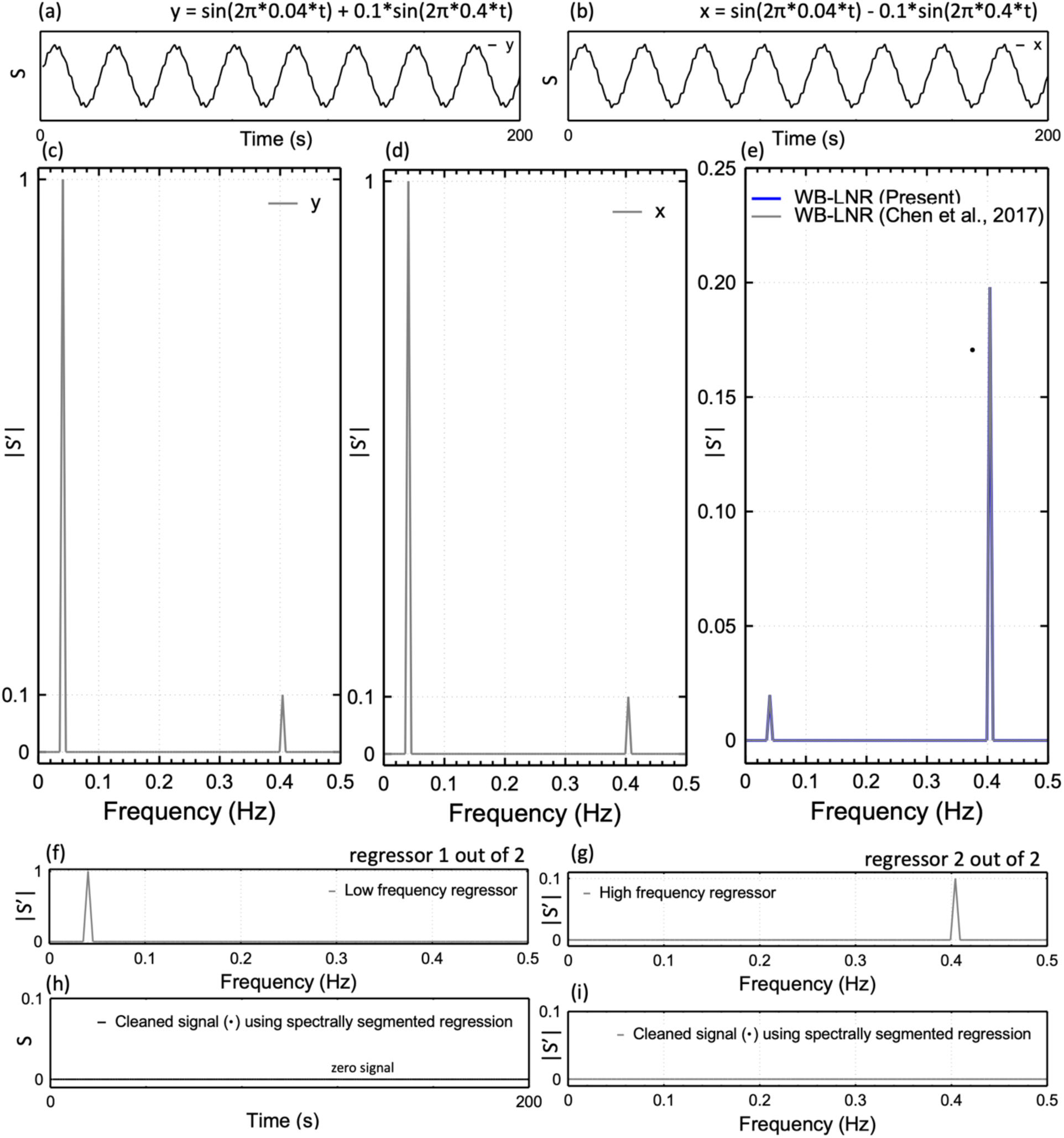
Comparison of whole-band linear regression (WB-LNR) and spectrally segmented regression for the simple case described in Figure 1 of Chen et al. [23] where (a) measurement y and (b) nuisance regressor x both contain low (0.04 Hz) and high (0.4 Hz)-frequency components, where the latter are mismatched. The corresponding magnitude spectra are shown in (c,d). (e) Whole-band linear regression (blue) leaves a significant residual, confirming the result from Chen et al. (black) (f,g) Low and high frequency regressors and (h,i) corresponding spectrally regressed time course and spectrum.

In a second step, we applied the method to a low-pass filtered resting-state signal time course, which was derived from an actual in-vivo resting-state signal time course by low-pass filtering to which different signal confounds were added (**Figs. 3,4**). These confounds consisted either of boxcars or of Gaussian noise using spectrally varying mixing coefficients (4 uniform segments, 0.2-0.5 mixing coefficients in 0.1 increments). Regression vectors were either the original boxcars or the original Gaussian noise. Regression was performed without and with regression of filtering residuals.

**Figure 3:**
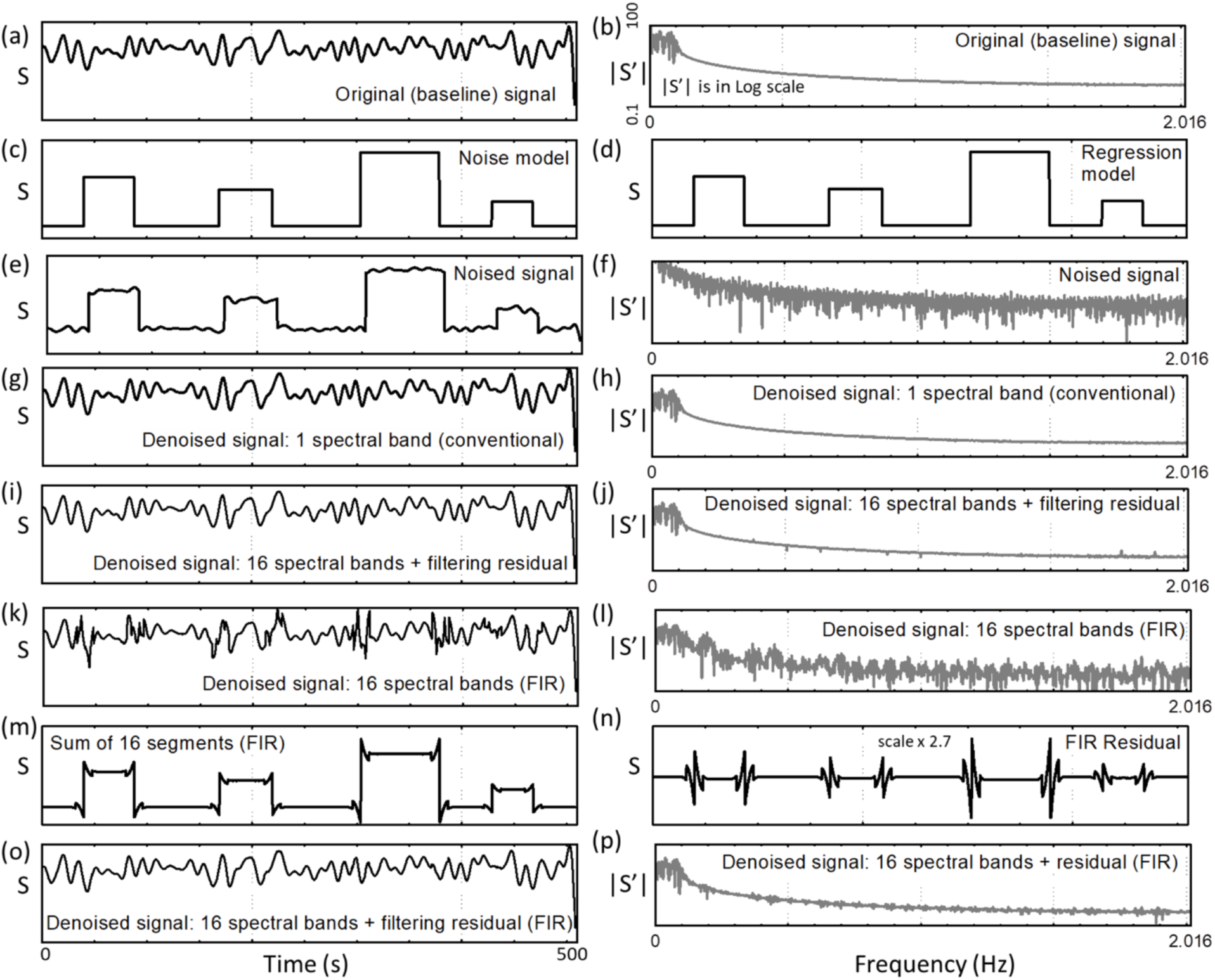
Ideal case for conventional regression where the regressor identically matches the nuisance component in the signal throughout the entire spectrum. The panels show (a) the original baseline signal in the time domain, (b) the signal in the frequency domain, the noise model, (d) regression model which matches the noise, (e-f) the noised signal before regression in the time domain and frequency domain, (g-h) the resulting signal from conventional regression which recovers the original signal, (i-j) the resulting signal from spectrally segmented regression with non-causal filtering, (k-l) the resulting signal from spectrally segmented regression with FIR segmentation without residual regression, and (m-p) the inclusion of the filtering residual and its effect on the regression. The sampling interval was 248 ms. The spectral-domain representation employs a logarithmic scale to enhance the visibility of signal differences at low powers.

**Figure 4:**
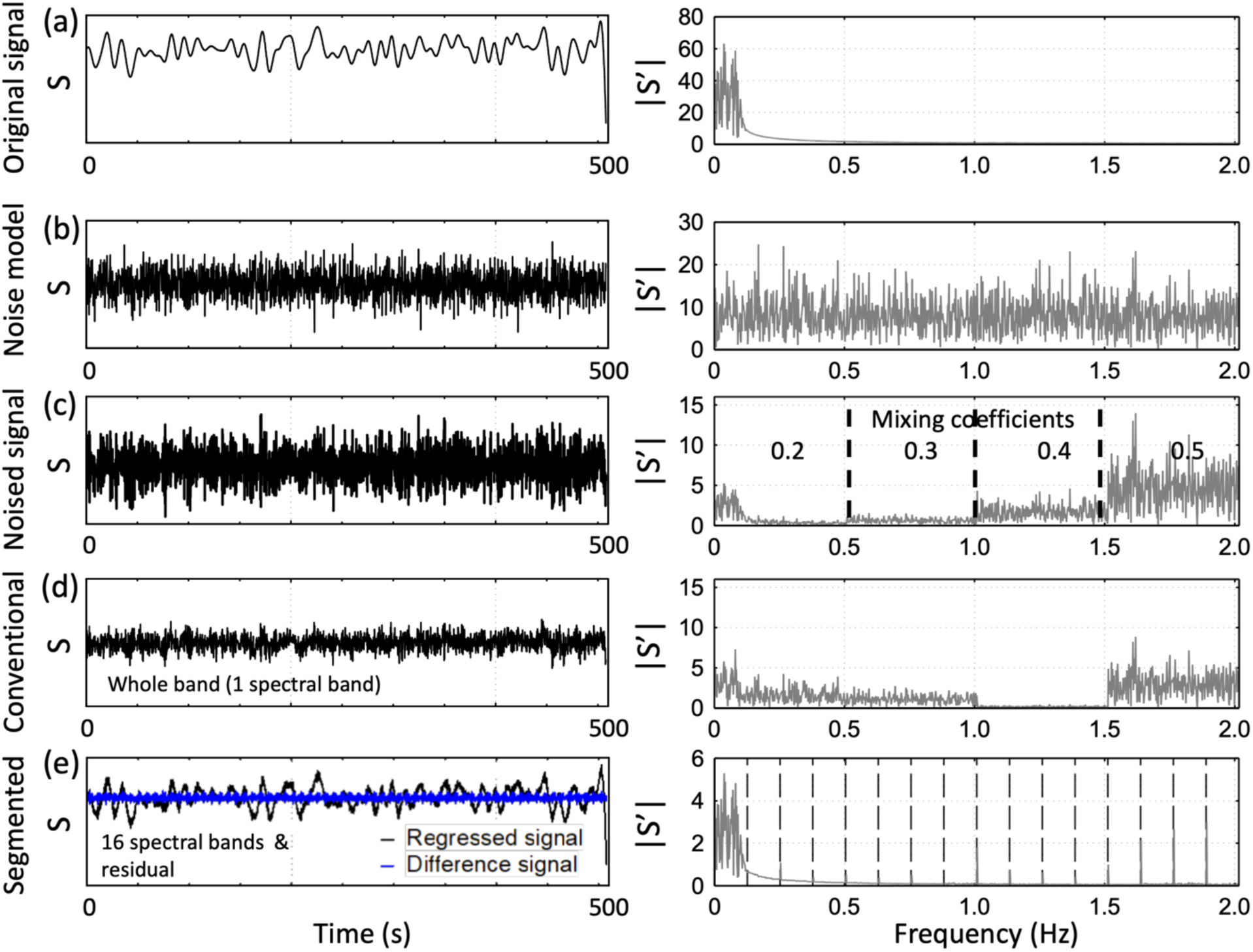
Simulation of a resting-state signal with confounding signal comparing whole band regression and spectrally segmented regression for the case of errors in the regression model. This figure demonstrates the ability of spectrally segmented regression to apply different regression weights to different spectral bands in the data, leading to efficient regression when uncertainties are present in part of the spectral domain. (a) Low-frequency resting-state signal. (b) Gaussian noise model used for regression. The (c) confounding signal was generated from (b) using spectrally varying mixing coefficients to create a mismatch (4 uniform segments, 0.2-0.5 mixing coefficients in 0.1 increments). (d) Whole band regression does not remove confounding noise. (e) Spectrally segmented regression using 16 spectral bands recovers the resting-state signal. The difference compared to the baseline signal is shown in blue.

In a third step, we simulated a low frequency and a high frequency two-node resting-state network in two brain slices (**Figs. 5,6**). Each slice consisted of 64×64 pixels with Rician noise signal time courses s(r) with an intensity range from 0 to 5 and 3000 time points with 0.205 sec time intervals, resulting in a frequency bandwidth of 2.43 Hz. Two circular seed regions were defined in left and right posterior brain regions (52 and 61 voxels, respectively) (Fig. 5a) and correlated at frequencies < 0.3 Hz (Fig. 5.b). Two circular seed regions were defined in left and right anterior brain regions (57 and 52 voxels, respectively) (Fig. 5a) and correlated at frequencies > 0.3 Hz (Fig. 5c). Correlations were created by mixing the signals s(r) in each pixels within these seeds with a Rician noise vector η that was either high pass filtered (> 0.3 Hz) or low pass filtered (< 0.3 Hz) with a mixing coefficient, *β*, of 0.3 according to Equation 8 [32].

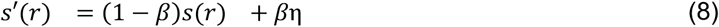

**Figure 5:**
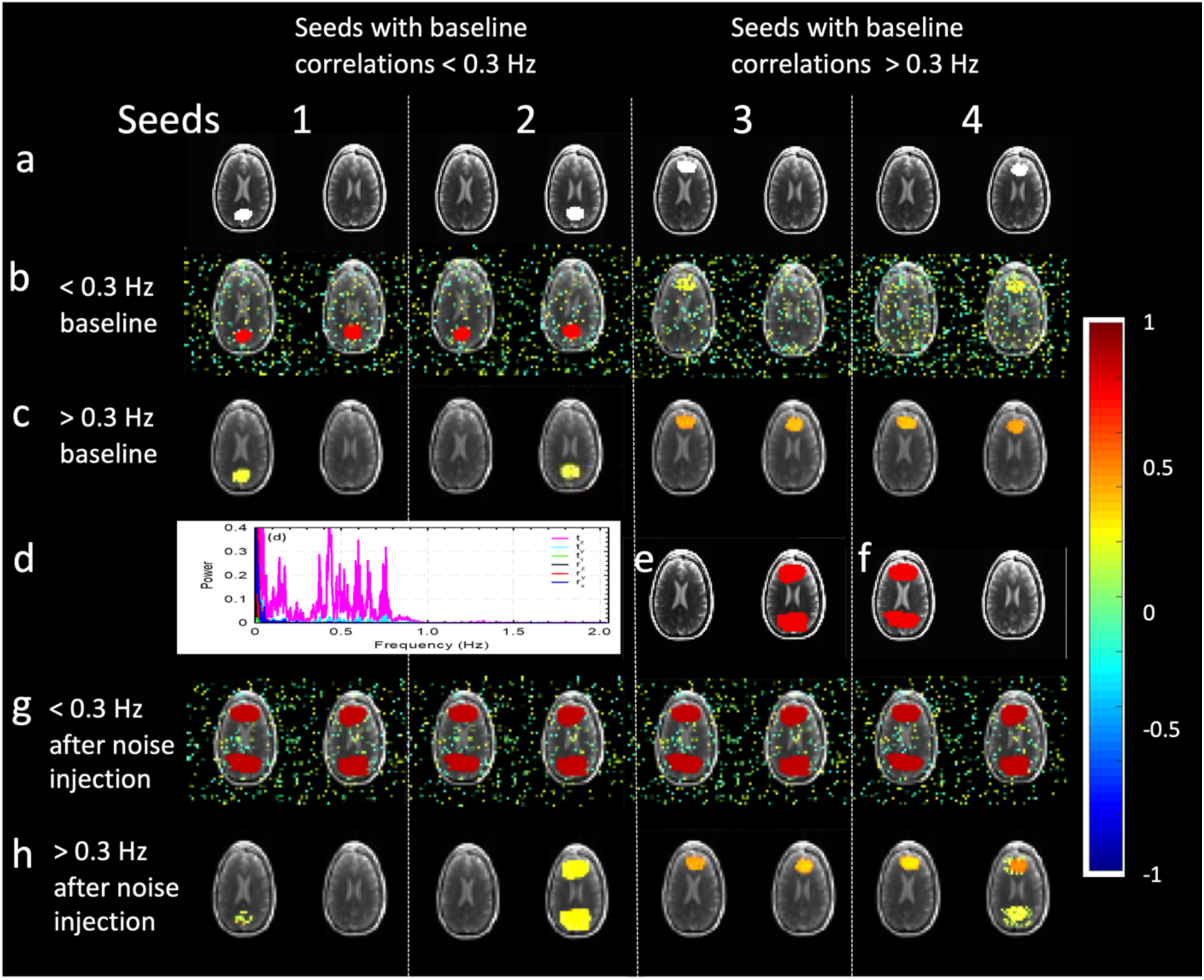
Simulation of a low frequency and a high frequency resting-state network in 2 brain slices before and after injection of low frequency and whole bandwidth noise. (a) Posterior seed regions (white) with baseline correlations < 0.3 Hz and anterior seed regions (white) with baseline correlations > 0.3 Hz. Corresponding seed-based correlations (b) at low frequencies < 0.3 Hz and (c) at high frequencies > 0.3 Hz. (d) Motion parameters from actual in-vivo scan, which were injected as whole band confounding noise into (e) the red noise regions in the right slice and after low-pass filtering (< 0.3 Hz) into (f) the red noise regions in the left slice. Note that the noise injection regions in e and f are larger than the original seeds 1-4. This is intentional as it simulates the injection of noise into correlated regions (seeds 1-4) and uncorrelated background regions surrounding the seeds, which contain Rician noise without explicitly defined correlations in the baseline model. Each seed simulates 50-60 signals in individual voxels while surrounding noise seeds simulate ∼85 signals in individual voxels. (g) Seed-based correlations after noise injection (h) at low frequencies < 0.3 Hz and (f) at high frequencies > 0.3 Hz. Correlation threshold: 0.1.

**Figure 6:**
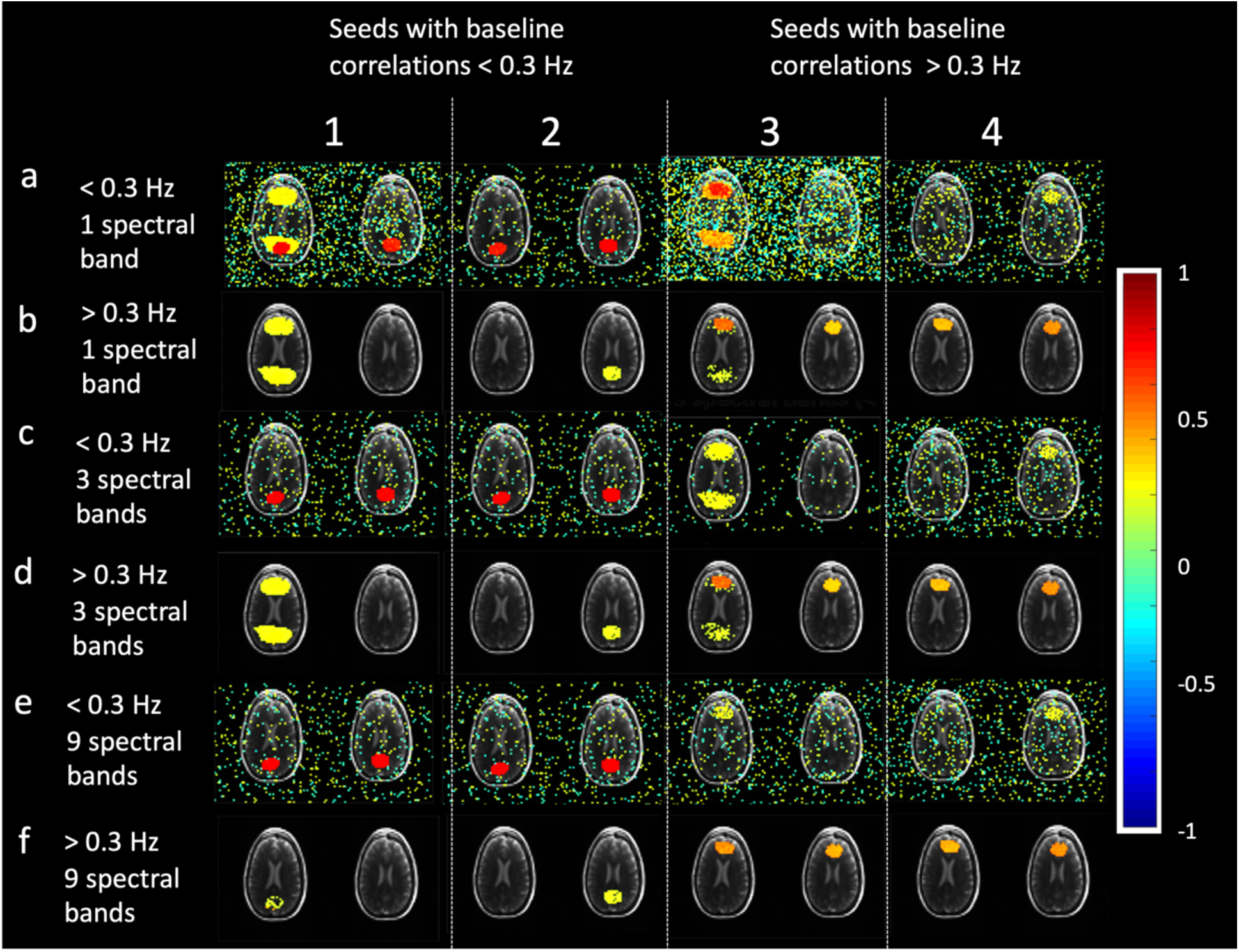
Simulation of denoising the low frequency (< 0.3 Hz) and high frequency (> 0.3 Hz) resting-state networks shown in Figure 5 after noise injection using spectrally segmented regression with different number of spectral bands of the regression vectors. (a,b) The simulations using a single spectral band demonstrate the failure of the conventional approach and the injection of artifactual connectivity in the high frequency regime (> 0.3 Hz) in the left slice. In the low frequency regime (< 0.3 Hz), conventional regression only partially succeeds and fails to remove the artifactual correlations in the noise regions surrounding the seed. (c,d) Spectrally segmented regression using 3 spectral bands of 0.81 Hz bandwidth shows reductions in regression errors. (e,f) Spectrally segmented regression with a sufficient number of bands (9 in this case with 0.27 Hz bandwidth) successfully recovers the original signals (Figure 5b,c) without injecting artifactual correlations. A minor, but observable, increase in correlations between Seed 3 and the background (Rician noise) above the correlation threshold (0.1) is noted for the conventional approach.

Whole frequency range noise (up to 2.43 Hz) based on motion translation parameters (tx, ty, and tz) parameters from an actual fMRI scan (Fig. 5d) was injected in the right slice into both seeds and into rim regions (85 voxels) surrounding these seeds that contain uncorrelated Rician noise (Fig. 5e), using mixing coefficients of 0.3, 0.4, and 0.5 for tx, ty, and tz, respectively. Low-pass filtered (< 0.3 Hz) noise based on these translation parameters was injected in the left slice into both seeds and into rim regions (85 voxels) surrounding these seeds that contain uncorrelated Rician noise (Fig. 5f), using the same mixing coefficients as in the right slice. This created a mismatch in the left slice between the injected noise and the regression vectors that were constructed from spectral segmentation of the original non-filtered translation parameters using non-causal filters. The rim regions allowed to assess the effect of regression on noise-injected pixels without underlying baseline resting-state correlations. The number of spectral segments was varied from 1 to 9 to investigate the effect of spectral segmentation on regression. Seed-based signal correlations with all pixels in the two slices were computed without regression, with whole-band linear regression (WB-LNR) and with frequency segmented regression.

Finally, the physiological noise regression approach in TurboFilt was compared to that of HRAN [31] using demonstration data provided with HRAN [https://github.com/LewisNeuro/HRAN] and default parameters provided with the HRAN demo script (**Fig. 7**). Spectrograms of the signals from the visual network were generated using the Chronux toolbox [http://chronux.org/].

**Figure 7:**
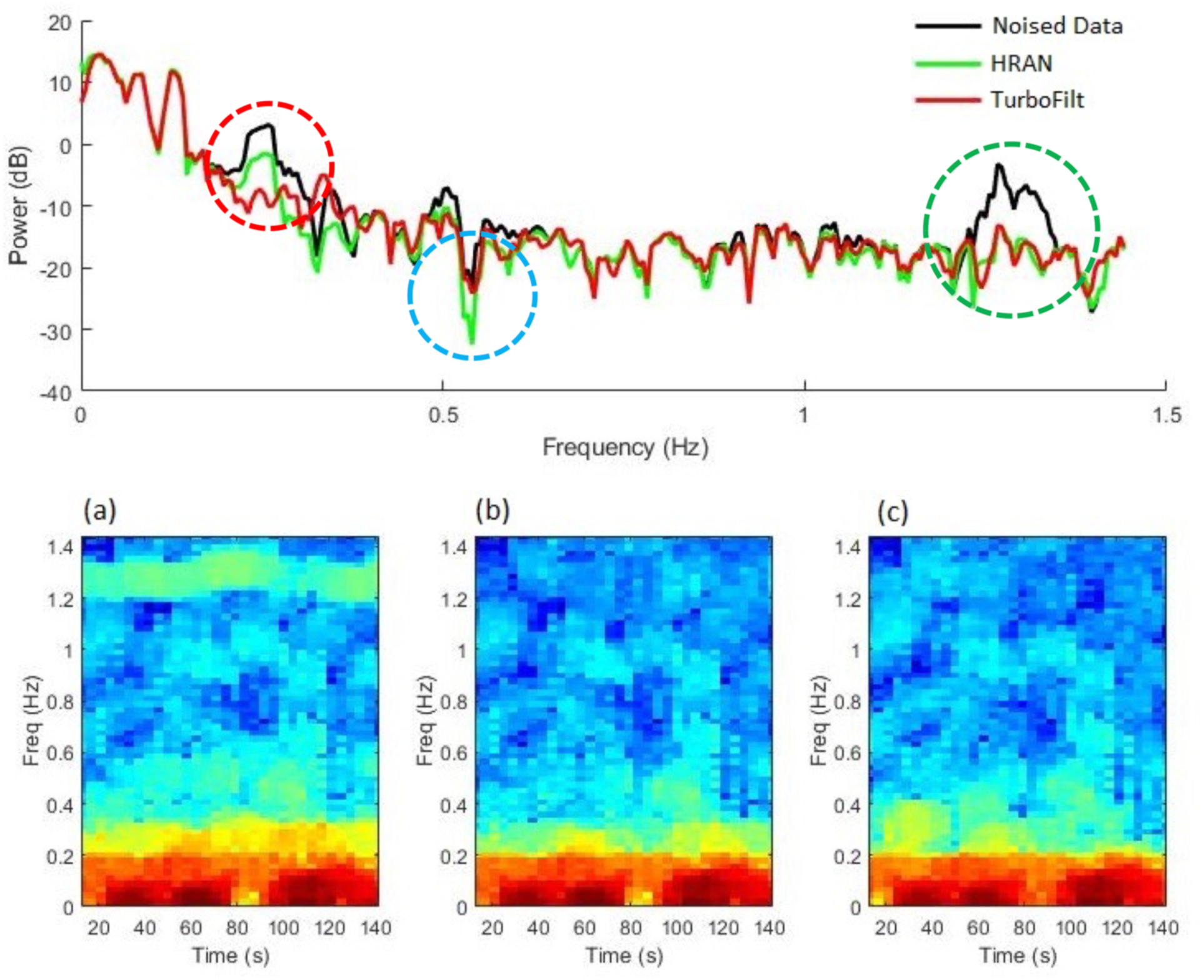
Comparison of physiological noise regression in HRAN and the spectrally and temporally segmented physiological noise regression using the demonstration data provided with HRAN acquired with a TR of 0.347 sec. The top figure shows spectra from a visual cortex seed before (Original Data) and after physiological noise regression using HRAN and spectrally and temporally segmented regression implemented in TurboFilt. Differences in the spectra are highlighted by dotted circles. Spectrograms are shown for (a) original data with physiological noise, (b) HRAN corrected data, (c) data corrected using spectrally and temporally segmented physiological noise regression.

### Subjects

Resting-state scans were acquired in five male right-handed healthy controls, a male patient with a low-grade glioma in insular, frontal and temporal cortex and a male patient with a glioblastoma in medial inferior temporal gyrus and temporo-occipital gyrus in the age range from 21 to 60 years. Subjects provided institutionally reviewed, written informed consent. Subjects were instructed to clear their mind and fixate on a cross-hair while keeping their eyes open. One of the healthy controls was scanned during capnometry controlled hyperventilation (p_ET_CO_2_: 19-25 mm Hg) to assess the robustness of the present method to increased and more variable levels of respiratory and cardiac signal pulsation.

### Data acquisition

Data were collected on a 3T Siemens TIM Trio scanner (Siemens Healthineers, Malvern, PA) equipped with MAGNETOM Avanto gradient system and 32-channel head array coil. High-resolution T_2_-weighted Turbo Spin Echo scans were acquired for anatomical reference. FMRI data were acquired in AC/PC orientation using (a) multi-slab echo-volumar imaging (MS-EVI) [14] (TR/TE: 246/25 ms, flip angle: 20°, no. of scans: 1000, 1100 or 1500, no. slabs: 4, spatial matrix per slab: 64×64×8, voxel size: 4×4×4 mm^3^, scan times: 4:10, 4:35 or 6:13 min; and (b) multi-band echo-planar imaging (MB-EPI) (https://www.cmrr.umn.edu/multiband/): TR/TE: 205/30 ms, flip angle: 15°, no. of scans: 3000, number of slices: 24, in-plane spatial matrix: 64×64, voxel size: 4×4×4 mm^3^, inter-slice gap: 0 mm, MB factor: 8, and scan time: 10:21 min. The cardiac and respiratory rates were recorded for physiological noise labeling.

### Data analysis

FMRI data were motion corrected and unilateral seed regions were manually selected in auditory, default mode, and visual networks in reference to the MNI atlas using TurboFIRE software [32]. Subsequent processing steps in TurboFilt included high-pass FIR filtering (0.005 Hz - 0.03 Hz), followed by either whole-band or spectrally and temporally segmented motion regression (12 uniform spectral segments using FIR filters and 8 temporal segments), and spectrally and temporally segmented physiological noise regression. FIR filters parameters were: 0.025 Hz transition bandwidth, Stopband Attenuation: 60 dB, Passband Ripple: 1 dB. White matter and CSF masks were manually generated to create physiological noise regression vectors. The physiological noise regression step was compared with HRAN (windows length 30 second, 50% overlap). Seed-based correlation analysis across the entire time series was performed with TurboFilt.

A second analysis pipeline performed spectrally segmented regression using TurboFilt and temporal segmentation using averaged sliding window correlation analysis (width: 15 s) with TurboFIRE software as described in [32] [33] to map the following networks: sensorimotor (SMN) – BA1-3, default mode (DMN) – BA7&31, visual (VSN) – BA17, and auditory (AUN) – BA41,42. Raw data were smoothed using an isotropic 5 mm gaussian filter.

To assess the convergence and stability of the approach when using a large number of spectral bands, high frequency (> 0.2 Hz) correlations in the sensorimotor resting-state network in the MS-EVI data set acquired in the healthy control during hyperventilation were mapped as a function of the number of spectral bands (1-64) using TurboFIRE with a 1 s sliding window.

## RESULTS

### Simulations

For the simple case described in Chen et al. [23], we observed the injection of artifacts into the high frequency component at 0.4 Hz with whole band linear regression (**Fig. 2e**) and incomplete removal of the low frequency nuisance component. Meanwhile, spectrally segmented regression using a low frequency regressor (< 0.25 Hz) (**Fig. 2f**) and a high frequency regressor (> 0.25 Hz) (**Fig. 2g**) mitigated the injection of artifacts and removed the low and high frequency nuisance components (**Fig. 2h and Fig. 2i**).

Computer simulations of regression of confounding structured noise signals show that spectrally and temporally segmented regression with inclusion of residual regression has comparable performance to conventional regression when the regression model is identical to the injected noise (**Fig. 3**). Performance degrades when regression of residuals is omitted, in particular for the case of FIR bandpass filters (**Fig. 3k,l**). Frequency-dependent errors in the regression model (**Fig. 4**) result, as expected, in failure of conventional regression (**Fig. 4d**), whereas spectrally and temporally segmented regression recovers the original signal (**Fig. 4e**). This was the case both for gaussian noise (**Fig. 4**) and for structured noise (results not shown).

Computer simulations of correlations in the resting-state network model at low (< 0.3 Hz) frequencies in posterior brain regions and high (> 0.3 Hz) frequencies in anterior brain regions with additional rim regions for noise injection illustrate the performance of the frequency segmented regression approach for a larger number of scenarios (**Fig.5**,**6)**. After noise injection all seed regions exhibit low frequency correlations due to the shared noise < 0.3 Hz (**Fig. 5g**). At high frequencies > 0.3 Hz, the anterior left seed 3 correlates with the anterior right seed 4 as in the original noise-free model (**Fig. 5h**). The anterior right seed 4, however, correlates additionally with the posterior right seed 2 where whole frequency range noise was injected (**Fig. 5g**), since correlations were introduced at frequencies above 0.3 Hz. Positive correlations at low frequencies are also found in all rim regions surrounding the seeds, as well as at high frequencies in the rim regions surrounding the seeds in the right slice, as expected.

Applying frequency segmented regression to the resting-state network model with noise injection in **Fig. 5** demonstrates the ability to recover the original noise-free brain model at high and low frequencies (**Fig. 6**). The conventional approach using whole-band regression vectors (1 spectral band) successfully removes the noise in the low frequency band (**Fig. 6a**) and recovers the original correlations when the regression vector matches the injected noise, which is the case for the posterior seed 2 in the right slice. However, whole-band regression fails to recover the original signals in case of low-pass filtered noise injected into the seeds and rims regions in the left slice and introduces false-positive correlations at low frequencies as observed in the correlation maps for both seeds in the left slice (**Fig. 6a**) and false-positive high frequency correlations for the posterior seed in the left slice (**Fig. 6b**). When using an insufficient number of spectral segments, as in the case of 3 segments, artifactual correlations can still be observed at high frequencies (**Fig. 6d**) despite the improvement in denoising at low frequencies (**Fig. 6c**) compared to using whole-band regression. Using 9 uniform spectral bands (0.27 Hz each) reduces artifactual low frequency correlations with anterior seeds (**Fig. 6e**) and strongly reduces artifactual high frequency correlations with posterior seeds (**Fig. 6f**). Positive correlations in the rim regions surrounding the seeds are removed by spectrally segmented regression and the original uncorrelated background is recovered.

The comparison of spectrally and temporally segmented physiological noise regression with HRAN (**Fig. 7)** using demonstration data from a visual cortex seed with narrow frequency range provided with HRAN shows that both approaches have similar performance. The spectra also show that HRAN and TurboFilt do not appreciably alter data outside the specified physiological noise search ranges for HRAN or labels in different segments in case of TurboFilt.

### In-vivo experiments

**Figs. 8 and 9** demonstrate the performance of the developed approach for denoising of high-speed fMRI data that were collected while the subject was hyperventilating, which introduced motion and physiological pulsation artifacts due to strong and rapid movement that covers a wide spectral range up to 0.75 Hz. The spectrally and temporally segmented motion regression step using 0.125 Hz spectral bandwidth substantially denoises signal components < 0.2 Hz in the visual network and CSF (**Fig. 8.b** and **Fig. 8.f**). Further reduction of respiration or cardiac signals was obtained with the physiological noise regression step in TurboFilt and HRAN (**Fig. 8.c** and **Fig 8.g**). Physiological noise regression in TurboFilt provides improved suppression of respiratory signal changes compared with HRAN (**Fig. 8.d** and **Fig. 8.h**). The frequency spectrum of the denoised signals (see the embedded figures in **Fig. 8.d** and **Fig. 8.h**) reflects the expected hemodynamic response models of the BOLD effect (e.g. [35, 36], [37]). **Figure 9** demonstrates that artifacts in the detection of low (<0.3 Hz) and high frequency (> 0.3 Hz) correlations in auditory, default mode, and visual networks at the edges of the brain and in white matter were considerably reduced when using spectrally and temporally segmented motion regression (**Fig. 9c,e**) compared to whole-band regression (**Fig. 9b,d**).

**Figure 8:**
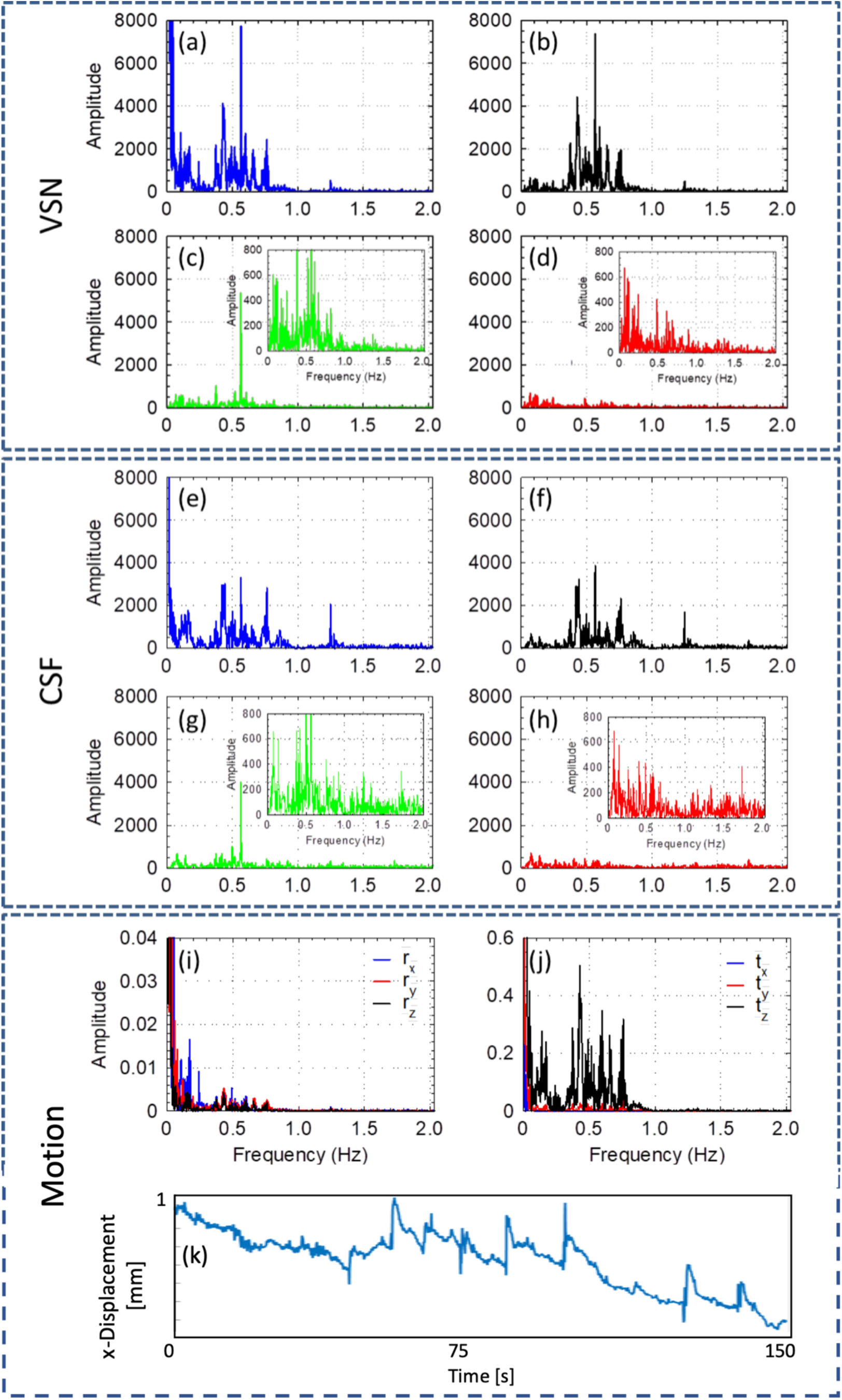
Different denoising steps to remove motion and physiological pulsation artifacts from (a-d) an occipital seed region in the visual network (VSN) and (e-h) a manually delineated region in cerebrospinal fluid (CSF) in a healthy control during a hyperventilation task. (i,j) The measured translation and rotation spectra show considerable spectral components up to 0.75 Hz. (k) Example of a motion parameter time course. (a.e) Raw data without denoising, (b,f) Spectrally and temporally segmented motion regression using TurboFilt (0.17 Hz spectral bandwidth), (c,g) Spectrally and temporally segmented motion regression using TurboFilt and physiological noise regression using HRAN, (d,h) Spectrally and temporally segmented motion regression and physiological noise regression using TurboFilt. MS-EVI data were acquired with TR of 246 ms, 32 slices and a scan time of 4:35 min.

**Figure 9:**
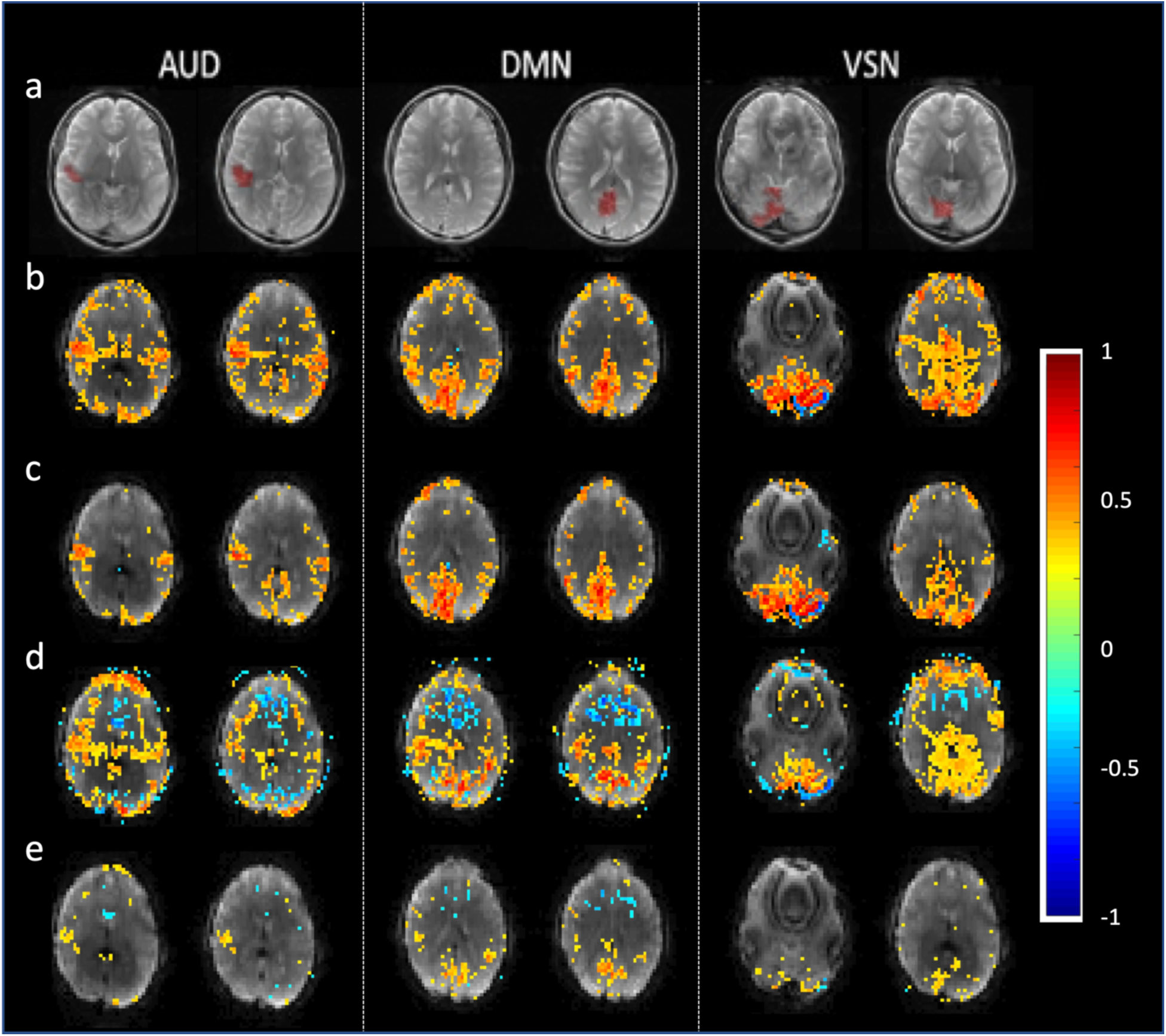
Low and high frequency signal correlations in three resting-state networks in a healthy control during a hyperventilation task, comparing (b,c) conventional whole band motion regression and (d,e) spectrally and temporally segmented motion regression (0.17 Hz spectral bandwidth) in two selected slices. (a) Seed regions in the auditory network (AUN), the default mode network (DMN), and the visual network (VSN) overlaid on T_2_-TSE scan. (b,c) Low frequency (< 0.3 Hz) correlations with (b) whole band motion regression and (c) spectrally and temporally segmented motion regression. (d,e) High frequency (> 0.3 Hz) correlations with (d) whole band motion regression and (e) spectrally and temporally segmented motion regression. Spectrally and temporally segmented motion regression reduces artifactual correlations along the periphery of the brain and in white matter, both at low and high frequencies. MS-EVI data were acquired with TR of 246 ms, 32 slices and a scan time of 4:35 min. Correlation thresholds: 0.3 for (b,c) and 0.25 for (d,e).

Spectrally and temporally segmented regression of motion and physiological noise using 0.125 Hz spectral bandwidth in combination with averaged sliding window correlation analysis further enhanced the detection of high-frequency (> 0.3 Hz) correlations in the auditory, default mode and sensorimotor networks (**Fig. 10.d**), which was co-localized with low frequency correlations below 0.3 Hz (**Fig. 10.b**). Spectrally and temporally segmented regression of physiological noise (**Fig. 10.d**) reduced artifactual correlations compared with HRAN (**Fig. 10.c**). Correlations at high frequencies > 0.3 Hz were also detected in the patients with brain tumors. **Figure 10** also shows an example of high-frequency correlations in the auditory network adjacent to a low-grade glioma in insular, frontal and temporal cortex that is posteriorly displaced and co-localized with low frequency (< 0.3 Hz) correlations. Similar to the healthy control, the suppression of physiological noise correlated with CSF pulsation at high frequencies > 0.3 Hz using spectrally and temporally segmented regression was stronger than with HRAN.

**Figure 10:**
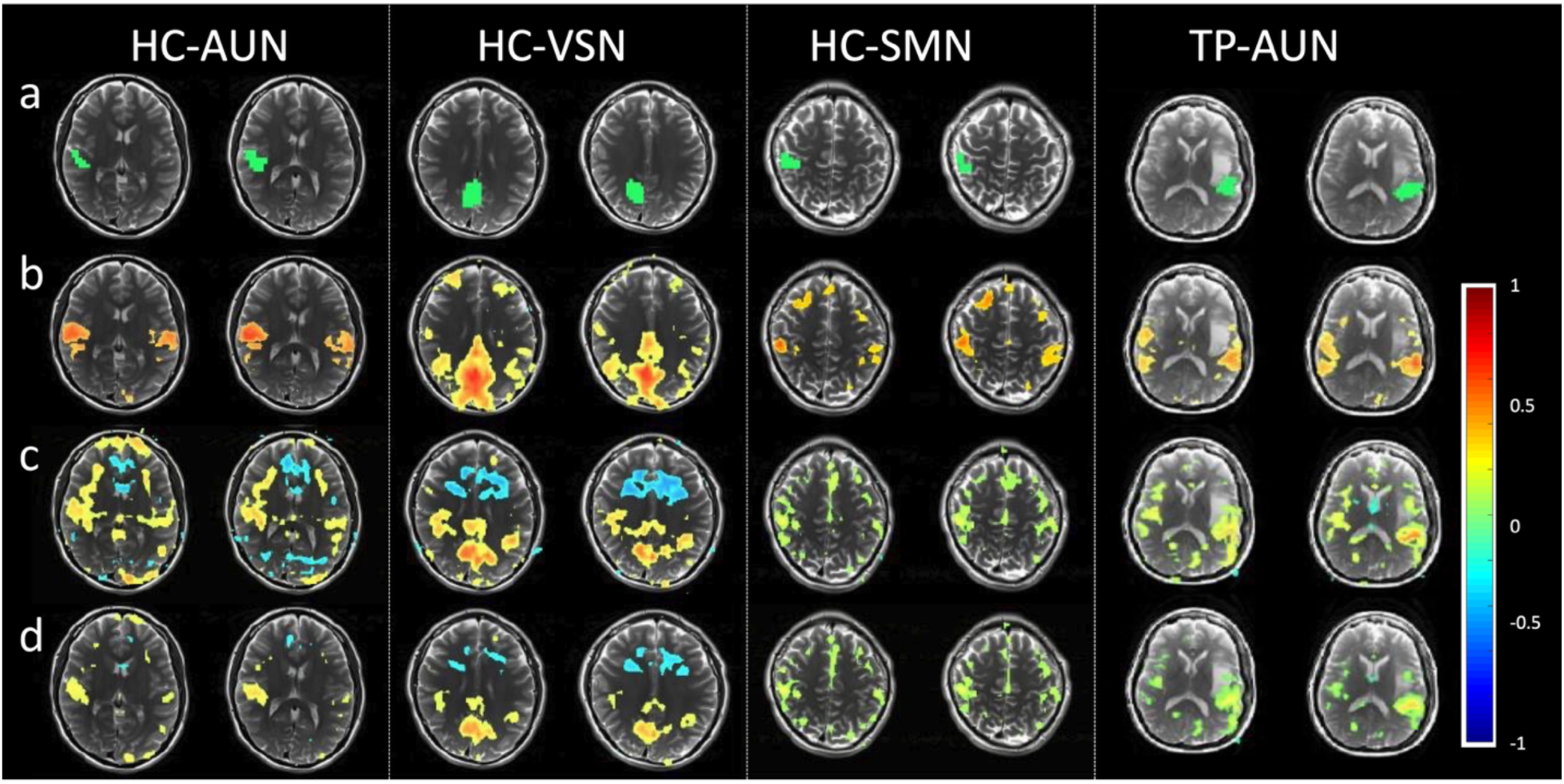
Low and high frequency signal correlations in three resting-state networks of a healthy control (HC) and in the peritumoral auditory network of a brain tumor patient (TP) using spectrally and temporally segmented motion regression and physiological noise regression (0.17 Hz spectral bandwidth) combined with averaged sliding window correlation for enhanced confound suppression. Unilateral (a) auditory network (AUN), visual network (VSN) and sensory motor network (SMN) seed regions overlaid on T_2_-TSE scan. (b) Low-frequency (< 0.3 Hz) correlation maps. (c,d) High frequency (>0.3Hz) correlation maps using (c) HRAN for physiological noise regression and (d) temporally and spectrally constrained regression of physiological noise showing stronger reduction of artifactual correlations along the periphery of the brain and in white matter. The healthy control data were acquired using MS-EVI with TR of 246 ms and scan time of 4:35 min. Correlation thresholds: 0.6 at low frequencies, 0.3 for high-frequency AUN and DMN maps and 0.2 for high-frequency SMN maps. The patient data were acquired using MB-EPI with TR: 205 ms and a scan time of 10:21 min. Correlation thresholds: 0.4 at low frequencies and 0.2 at high-frequencies.

False-positive high frequency correlations in cortical areas and white matter regions decreased asymptotically with increasing number of spectral bands while the sensitivity for detecting true positive correlations decreased only slightly with increasing number of spectral bands. An example data set acquired during hyperventilation that was analyzed with up to 64 spectral bands to map sensorimotor correlations is shown in the **Figure S1.** There was a considerable decrease of artifactual correlations outside of the sensorimotor cortex from 1 to 4 spectral bands (0.5-2 Hz spectral bandwidth), in particular in inferior brain regions. Further moderate improvements were observed when increasing the number of spectral bands to 16 (0.125 Hz spectral bandwidth), and no significant improvement were observed between 16 to 64 spectral bands but the results remained stable. The sensitivity for detecting true positive correlations in sensorimotor cortex decreased only slightly with increasing number of spectral bands and asymptotically reached a stable correlations pattern when using 4 or more spectral bands.

## DISCUSSION

The present study demonstrates that spectral and temporal segmentation of motion parameters and physiological noise signals represents a powerful and novel regression approach for minimizing confounding non-neuronal signal fluctuations in resting-state fMRI data. As our results demonstrate, the approach is particularly powerful for removing movement and respiration related signal changes that cover extended frequency ranges that cannot be approximated by single frequency regression vectors.

Our simulations show that original signals and correlations are recovered when a sufficient number of spectral bands is used. For the simulations in **Figure 6**, ideal regression can also be achieved by segmenting the regression vectors at 0.3 Hz. However, this requires that the spectral band of interest for connectivity analysis to be defined prior to denoising, which limits the usefulness of the approach for situations where the frequency spectrum of the correlations of interest are not known a priori. Additionally, the performance of the denoising process will be undermined if inaccuracies are present in a spectral segment of the regression vector within the spectral band of interest. Use of a sufficiently large number of uniform spectral bands mitigates these problems.

It is, therefore, important to understand whether there is an upper limit on the number of uniform spectral segments that could be used. The in vivo data suggest that a regressor spectral bandwidth between 0.125 and 0.5 Hz depending on the motion spectral pattern is effective for motion regression. Using an excessively large number of spectral segments can weakly compromise the effectiveness of the regression process especially when the regression vectors can be measured with high accuracy and increases the impact of regression errors at the edges of the spectral bands on overall regression performance. A further limitation is given by the digitization of frequency spectra. On the other hand, the results in **Figure 6** show that increasing the number of segments is beneficial when inaccuracies are present in part of the spectral domain of the regression vectors. Therefore, the recommendation is to use the maximum number of uniform spectral bands constrained by the loss of degrees of freedom and the tolerated level of residual correlations due to spectral segmentation and by spectral digitization. Our observations are consistent with the analyses by Chen et al [23]. However, it was found in the present study that the artifactual networks observed in high frequencies do not necessarily represent low frequency connectivity, but rather regions with incompletely sampled intra-scan motion.

When combined with temporal segmentation, the number of spectral bands may be additionally limited by the length of the temporal window in order for the system of equations not to be overdetermined. For this reason, phase shifting of regression vectors in physiological noise regression is preferred to increasing the number of regression vectors using a fixed array of phase shifts, both of which gave nearly similar results. An alternative approach for temporal segmentation is the sliding window approach [32] that provides powerful rejection of transients.

As physiological noise is estimated from motion regressed data obtained after spectrally and temporally segmented regression of motion parameters, there will be no shared information between the motion regressors and the physiological noise regressors. The physiological noise regressors effectively represent the residual physiological noise that was not removed by motion regression. An exception to this would be if large spectral bands that are much wider than the labeled physiological noise bands are used in motion regression. In that case, it is conceivable that information might be shared between the motion regressors and the physiological noise regressors estimated from the motion regressed data. Nevertheless, double regression of the same information in sequence will not introduce artifactual correlations as the calculated weights in the second regression approach zero.

Our simulations (**Figs. 6.b,d)** demonstrate that artifactual correlations is injected in high frequencies (> 0.3 Hz) when low-passed noise (< 0.3 Hz) that is mixed into the original signals is regressed using whole-band nuisance regressors that match low frequency components (< 0.3 Hz) of the injected noise but not high frequency components (> 0.3 Hz). Artifactual correlations are, therefore, understood to be injected into high frequencies when the nuisance regressors match the signals in the high-power, low-frequency part of the signal but not the high-frequency, low-power parts. Spectral segmentation mitigates this problem as nuisance in different spectral bands is represented by independent regression vectors that are assigned independent weights. The high-pass filter in conventional whole-band regression is in principle equivalent to a 2-band spectral segmentation with data below the high pass limit effectively being discarded. The present technique preserves data in the entire frequency spectrum and does not necessitate the use of high pass filtering and, therefore, mitigates making assumptions about the characteristics of the low frequency end of the underlying nuisance signals. It is particularly applicable to investigating high frequency correlations as a large number of bands can be used in nuisance regression allowing for unbiased nuisance regression at higher frequencies.

The technique is consistent in principle with the recommendations presented in the discussion of Chen et al [24] who recommended post-processing the data in spectral bands of interest. The present approach, however, does not spectrally isolate the data and relies on spectral segmentation of the regressors (e.g. motion parameters) without filtering the scan data or defining spectral bands of interest a priori, unlike the model-based HRAN technique [31] which does not regress whole-band noise to avoid the injection of artifactual correlations.

The in vivo results provide evidence for resting-state networks at frequencies above 0.3 Hz in both healthy controls and patients with brain tumors. Consistent with the results shown in[33] our present data confirm that spatial displacement of resting-state correlations due to a tumor at low and high frequencies is comparable, which supports the physiological origin of high frequency correlations.

### Limitations

The physiological noise regression approach described herein requires short enough TR times to obtain non-aliased representation of physiological noise which is necessary in the labeling step. Further, the bandwidth of the labeled physiological noise is assumed to be much narrower than that of the entire signal. If the data is acquired with a TR that is long enough to partly or fully undersample the typical physiological noise bandwidth, the present approach will not be applicable. Analyzing the connectivity pattern of the removed variance as described by Bright et al.[38, 39] would allow to determine the useful number of frequency bands. This is the subject of a future publication. An analytical or numerical approach to predict the performance of our approach as a function of the number of spectral bands is desirable but beyond the scope of the current study. The decrease of the effective degrees of freedom with increasing number of spectral segments and its impact on sensitivity require signal variance estimation using for example the xDF approach [40]. There is a need for formal statistical of the accuracy/complexity tradeoff of increasing the number of regressors in our approach, for example using the MACS toolbox [41]. These, however, are beyond the scope of the current study. Finally, digitization will limit the number of spectral bands, in particular at long TR, and the applicability of this approach to frequency segmented regression within short sliding windows.

The current implementation of the methodology in TurboFilt employs non-tapered sequential windows, in part due to computational constraints, which could introduce artificial jumps in the spectra of the regression vectors. An implementation of spectral segmentation of regression vectors in TurboFIRE, which provides quasi-continuous sliding-window correlation analysis with regression, is under development. This also addresses rippling effects due to the sinc convolution of the window and spill-over of signals into other time windows. Further studies are required to assess the limitations of tapering the window, which broadens the spectral features and may introduce regression errors.

Phase delays in the current implementation are performed on a slice-by-slice basis due to computational constraints. Patch-wise phase delays could be implemented to improve the performance of physiological noise regression within signal-to-noise constraints.

While our physiological noise regression step does not employ autoregressive modeling, frequency segmentation is expected to mitigate the possibility of autocorrelations biasing particular frequencies. The current approach using regression of residuals further reduces the impact of autocorrelations. It is feasible to regress residuals per frequency band, but the stability of this approach against overfitting remains to be characterized. A thorough investigation of the autoregressive order of the residuals is beyond the scope of the current study.

Manual labeling of physiological noise as performed in the current study could be automated using the peripheral recordings of heart rate and breathing with modeling of harmonics, which is expected to improve the robustness of our methodology.

The multi-band EPI sequence requires very high acceleration factors while the multi-slab EVI uses lower acceleration, which reduces aliasing, however at the expense of blurring in the slice dimension due to T_2_* relaxation. It is expected that tSNR improvements may differ for the two sequences and depend on the choice of reference partition as shown in[42, 43].

## CONCLUSIONS

This novel spectrally and temporally segmented regression approach considerably improves the suppression of motion and physiological noise in high-speed resting-state fMRI data compared with conventional whole band regression at both low and high frequencies and minimizes the introduction of spurious components into higher frequencies thereby improving the fundamental understanding of brain connectivity dynamics.

## Data Availability

All data produced in the present study are available upon reasonable request to the authors

## Acknowledgements

Supported by NIH grant 1R21EB022803-01. We gratefully acknowledge Essa Yacoub, Sudhir Ramanna and Steen Moeller for their contributions to the development of multi-slab and multi-band echo-volumar imaging. We gratefully acknowledge Dr. Arpad Zolyomi for supervising data collection and controlling the capnic state during respiratory challenge studies. We thank Kishore Vakamudi for help with subject recruitment, data acquisition and preliminary data analyses.

---

**Supporting Figure S1:**
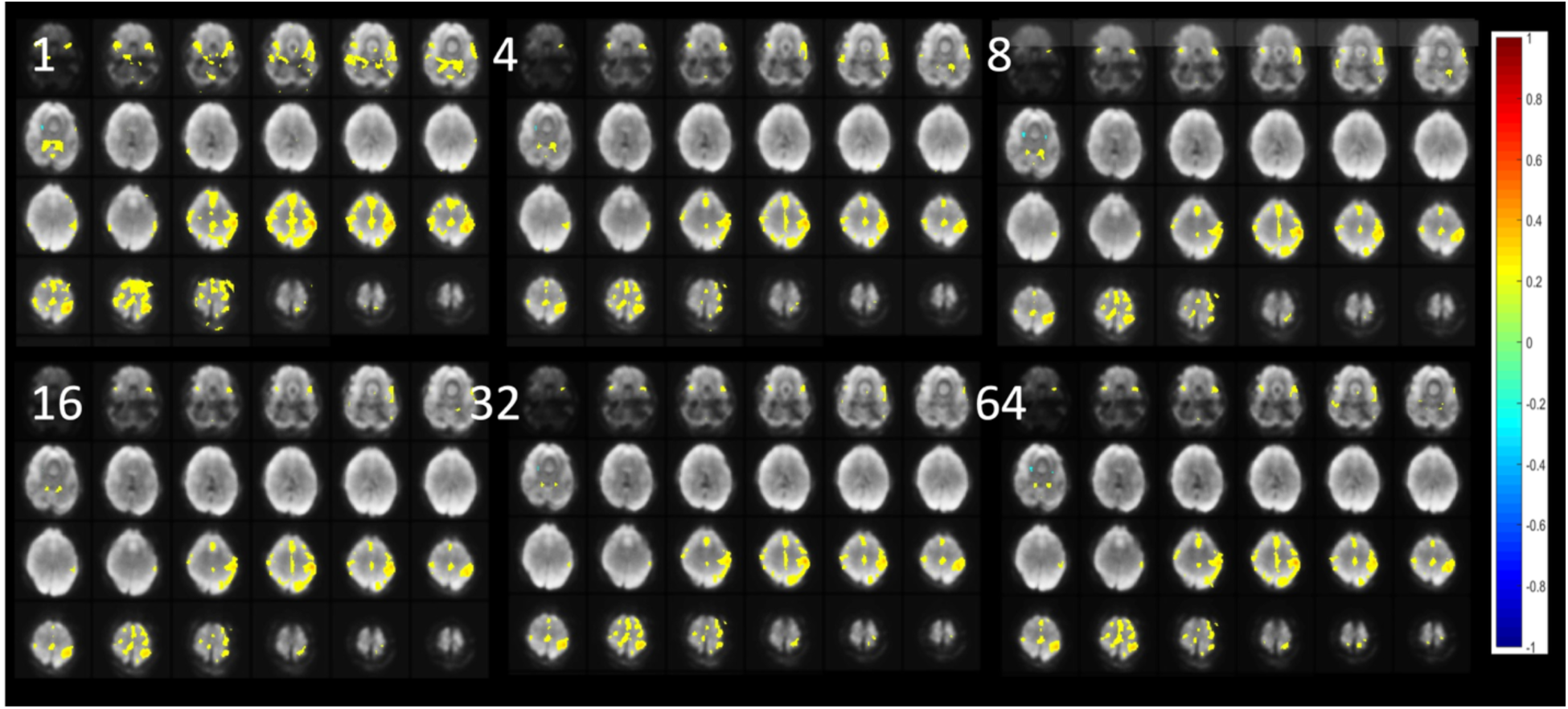
High frequency (> 0.2 Hz) sensorimotor correlations maps as a function of the number of spectral bands (1, 4, 8, 16, 32 and 64) using unilateral sensorimotor seed. MS-EVI data were acquired with TR of 246 ms, 32 slices and a scan time of 4:35 min. Averaged sliding window correlation analysis was performed using a 1 s window width and a correlation threshold of 0.3.

